# The 1st year of the COVID-19 epidemic in Estonia: an interrupted time series of population based nationwide cross-sectional studies

**DOI:** 10.1101/2021.09.06.21263154

**Authors:** Anneli Uusküla, Ruth Kalda, Mihkel Solvak, Mikk Jürisson, Meelis Käärik, Krista Fischer, Aime Keis, Uku Raudvere, Jaak Vilo, Hedi Peterson, Ene Käärik, Mait Metspalu, Tuuli Jürgenson, Lili Milani, Liis Kolberg, Ene-Margit Tiit, Kristjan Vassil

**Affiliations:** Institute of Family Medicine and Public Health, University of Tartu, Estonia; Johan Skytte Institute of Political Studies, University of Tartu, Estonia; Institute of Mathematics and Statistics, University of Tartu, Estonia; Institute of Computer Science, University of Tartu, Estonia; Institute of Genomics, University of Tartu, Estonia; Vice-Rector, University of Tartu, Estonia

**Keywords:** COVID-19, SARS-CoV-2, prevalence, surveillance, epidemiology, health inequalities, Estonia

## Abstract

**Background:** Decisions about the continued need for control measures and the effect of introducing COVID-19 vaccinations rely on accurate and population-based data on SARS-CoV-2 positivity and risk factors for testing positive.

**Methods:** In this interrupted time series of population-based nationwide cross-sectional studies, data from nasopharyngeal testing and questionnaires were used to estimate the SARS-CoV-2 RNA prevalence and factors associated with test positivity over the 1^st^ year of the COVID-19 epidemic.

The study is registered with the ISRCTN Registry, ISRCTN10182320.

**Results:** Between April 23, 2020 and February 2, 2021, results were available from 34,915 individuals and 27,870 samples from 11 consecutive studies. The percentage of people testing positive for SARS-CoV-2 decreased from 0.27% (95% CI 0.10% - 0.59%) in April to 0.04% (95% CI 0.00% - 0.22%) by the end of May and remained very low (0.01%, 95% CI 0.00% - 0.17%) until the end of August, followed by an increase since November (0.37%, 95% CI 0.18% - 0.68%) that escalated to 2.69% (95% CI 2.08% - 2.69%) in January 2021. In addition to substantial change in time, an increasing number of household members (for one additional OR 1.15, 95% CI 1.02-1.29), reporting current symptoms of COVID-19 (OR 2.21, 95% CI 1.59-3.09), and completing questionnaire in the Russian language (OR 1.85, 95% CI 1.15-2.99) were associated with increased odds for SARS-CoV-2 RNA positivity.

**Conclusions:** SARS-CoV-2 population prevalence needs to be carefully monitored as vaccine programmes are rolled out in order to inform containment decisions.

**Strengths and limitations of this study:**

- Our study relies upon nation-wide and population-based data on SARS-CoV-2 prevalence, and presents changes in prevalence over the whole 1st year of the Covid-19 epidemic.
- Our analysis of SARS-CoV-2 infection risk factors is not limited to notification or health care-based case data.
- Selection bias may have been introduced as a result of low response rate. The direction of bias is unclear, but most likely operates rather uniformly over the period of observation, though this presents less of a threat to the SARS-CoV-2 prevalence trend analysis.
- Our data could be used to adequately project the future course of the SARS-CoV-2 epidemic and the effect of control measures.

## Introduction

On March 11, 2020, the World Health Organization (WHO) characterised COVID-19 – a condition caused by the severe acute respiratory syndrome coronavirus 2 (SARS-CoV-2) – as a pandemic [1]. The ongoing pandemic of SARS-CoV-2 is following in the footsteps of the major pandemics of the past century (the influenza A virus pandemic in 1968 (H3N2) and 1918 (H1N1)). Within a year of the pandemic, the world has passed the milestone of two million COVID-19 deaths [2].

At the beginning of the 2^nd^ year of the epidemic, the highest daily new COVID-19 case notification rates (over 1,000 new daily cases per million inhabitants) have been reported from a small country in Europe – Estonia [3].

Control measures (nonpharmaceutical interventions, including business and school closures, restrictions on movement, and total lockdowns, social distancing) have been widely implemented to contain the spread of SARS-CoV-2 and have been effective in curbing the COVID-19 epidemic, but they do not represent desirable long-term strategies. The future trajectory of the COVID-19 pandemic hinges on the dynamics of both viral evolution and population immunity against SARS-CoV-2.

Understanding the future trajectory of this disease requires knowledge of the population-level landscape of immunity, generated by the life histories of the SARS-CoV-2 infection or vaccination among individual hosts [4]. The drivers of future COVID-19 dynamics are complex. However, characterisation of the pre-vaccination prevalence, the change of active infections, and development of immunity in the population are vital data elements for adequately projecting the future course of the SARS-CoV-2 epidemic and the effect of containment measures.

The evidence of the first year of the COVID-19 epidemic is frequently based on data from symptomatic patients,[5,6] seroepidemiological studies,[7] and modelling [8]. Most studies are based on small or selected population samples (e.g. hospital admissions) providing data not representative of the community. To the best of our knowledge, large population-based studies needed to understand risk factors, dynamics and delineating the pre-vaccination course of the COVID-19 pandemic, are scarce [9,10].

In this study, we rely on a national survey designed to be representative of the target population to describe the course of the epidemic over the first year and risk factors for testing positive for SARS-CoV-2 in Estonia (until the end of January 2021).

## Background

### National testing and case notification data

In Estonia, as of January 31, 2021, 785,333 SARS-CoV-2 (RNA) tests (58,967 per 100,000 population) with a cumulative test positivity rate of 6.1% (range 0-29%) had been undertaken for 472,356 unique individuals (Figure 1). The test positivity rate was highest at the beginning of the epidemic (29% on March 12, 2020), and with a daily average of 12.7% over December 2020 to January 2021.

**Figure 1.**
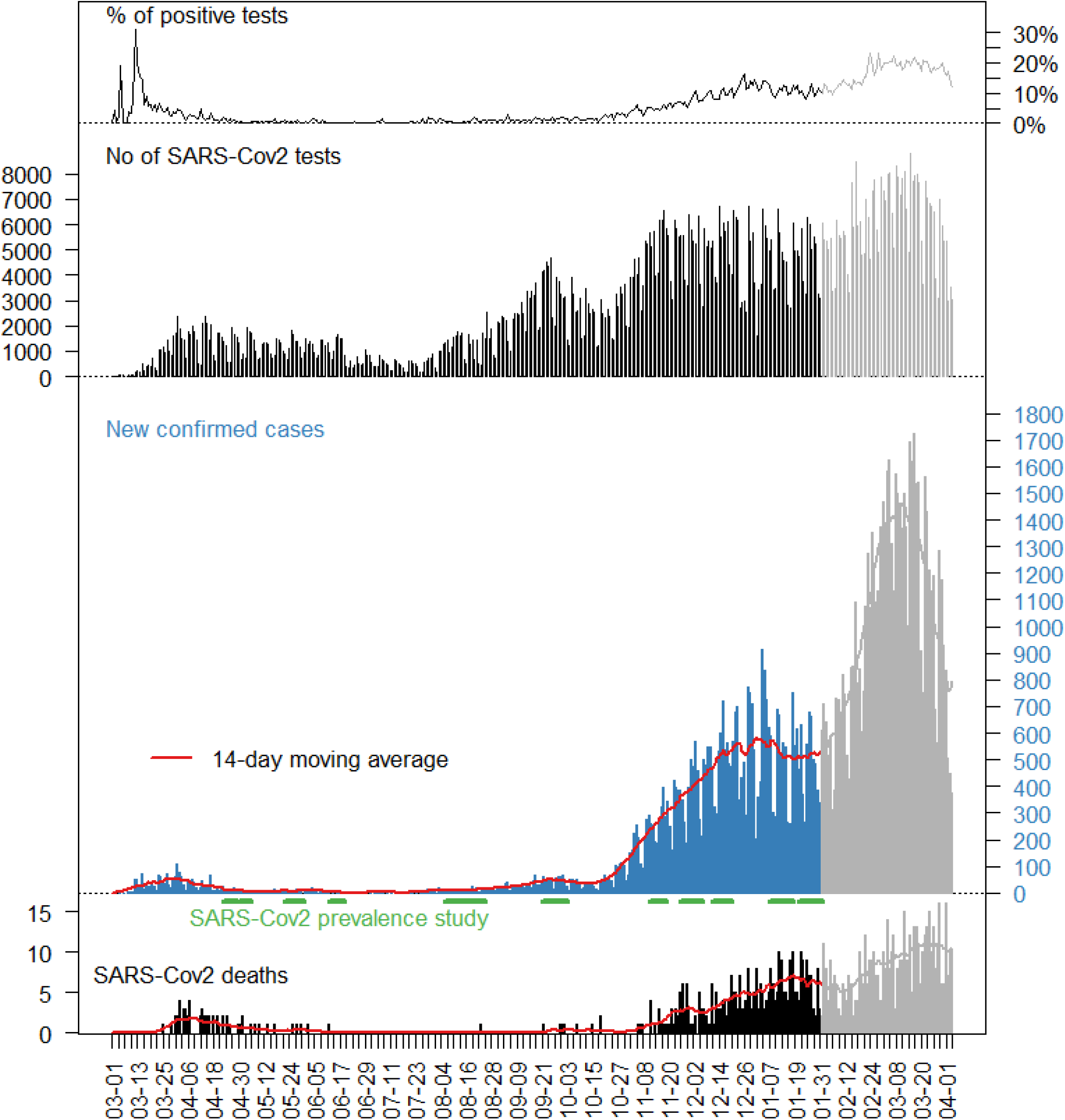
The COVID-19 epidemic in Estonia: daily numbers of new confirmed cases, number of tests, proportion of positive tests, and number of deaths, 2020-2021.

**Figure 2.**
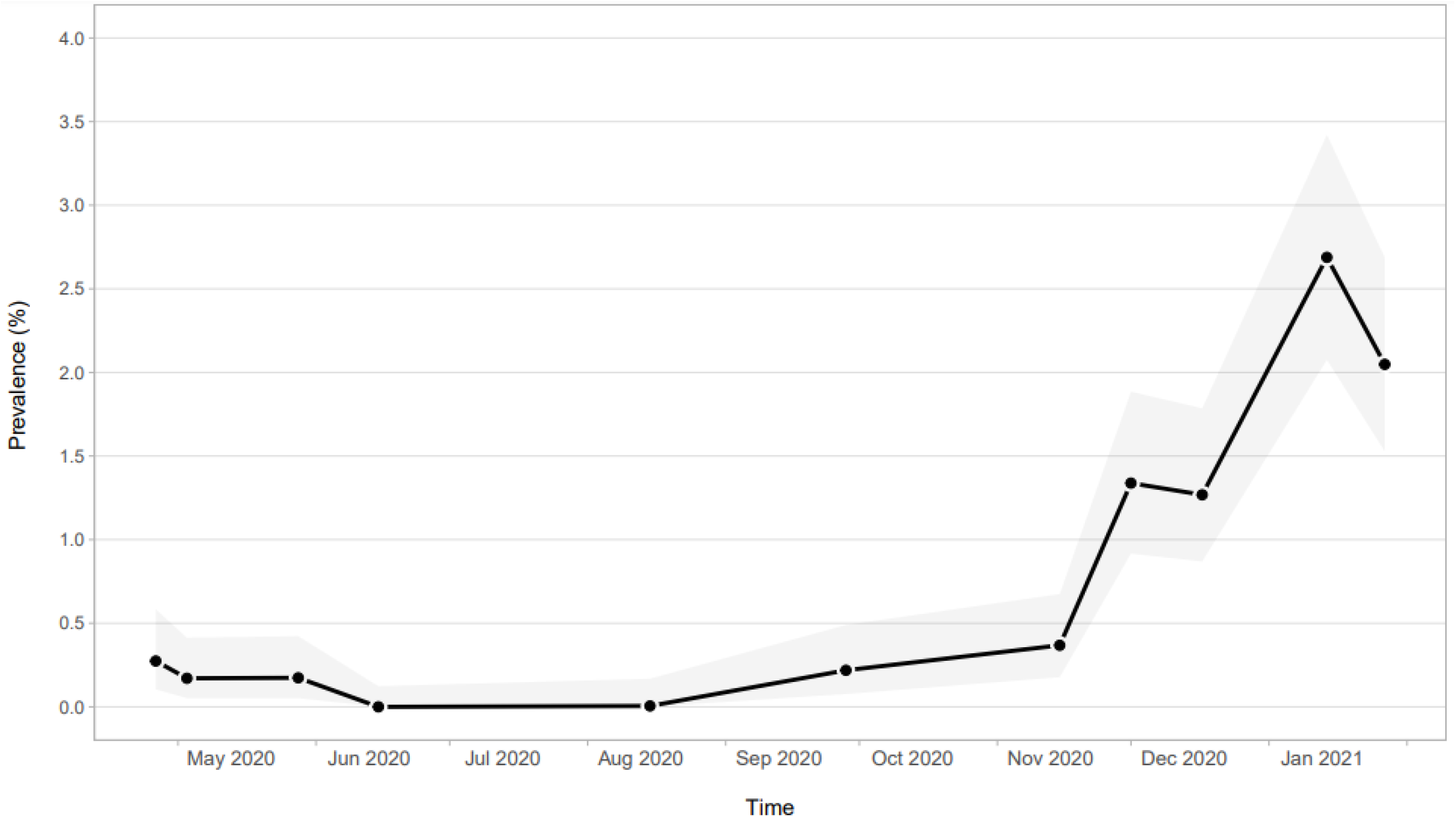
Percentage of population testing positive for SARS-CoV-2 over time in Estonia during the 1^st^ year of the COVID-19 epidemic 2020-2021. The grey area indicates 95% confidence intervals.

As of January 31, 2021, the total number of confirmed COVID-19 cases in the country was 44,208 (3,326 per 100,000). The highest number of new cases per day (*n*=1,101, 8.2 per 100,000) was reported on January 6, 2021 [12]. Of the confirmed cases, 55% were female, with case rates of 3,407 and 3,149 per 100,000 among men and women, respectively. The confirmed SARS-CoV-2 case rate was the highest among people aged 15-24 years (4,120/100,000), followed by the age group 45-54 years (4,053/100,000), and lowest among children aged less than 10 years (522 and 1,279 per 100,000 among 0-4 and 5-9 year olds, respectively).

By January 31, 2021, of all the confirmed COVID-19 cases, 5.6% (n=2,471) had been hospitalised for treatment and 0.9% (*n*=419) had died.

Since February 2021, Estonia has witnessed a constant increase in the daily number of new COVID-19 cases (with the highest 14-day cumulative number of cases per 100,000 of 1,553 on March 19, 2021) [3].

### Response to the epidemic

The first case of COVID-19 was confirmed in Estonia on February 26, 2020 [3]. A special digital referral system was developed in mid-March to simplify the referral process [12]. Individuals who were deemed to be at high risk for the SARS-CoV-2 infection (symptomatic patients referred by family physicians) and frontline staff (health care, nursing home, social workers, police, and border guard officers with a referral letter from their employer) were all eligible for testing. Testing eligibility was relaxed by July 2020.

On March 13, 2020, the Estonian government declared a state of emergency. The emergency measures did not include aggressive lockdown rules – people were allowed to leave their homes at any time so long as they observed social distancing. By June 2020, the restrictions were gradually eased but physical distancing requirements, i.e. the 2+2 rule (up to two people can be in a public place together and at least a 2-meter distance must be kept from others [13]), have remained in force. In response to the increase of new case notifications since the last week of July 2020, and attributing the new cases to visiting nightclubs and bars, the Police and Border Guard Board imposed bans on night-time alcohol sales from August 7 (in two counties) [14] and, since September 25, a nationwide restriction on the sale of alcohol has been in force. Since the beginning of November 2020, additional measures on the workplace (recommendation to work remotely, and cancelling all joint events), in public places and transport (mandatory mask wearing) were implemented [15].

COVID-19 vaccination started in January 2021 [16].

## Methods

### Source population

In 2020, the population of Estonia was estimated at 1,326,535 million people (equivalent to 0.02% of the total world population), with 68% of the population living in urban areas. The Estonian language is spoken by roughly 68% of the population, with approximately 28% of the population being Russian speakers [17]. Historically, most of Russians speaking population is settled in the capital, Tallinn, or the north-eastern region of the country (Ida-Virumaa county) [18].

### Data source - *SARS-CoV-2 community prevalence studies*

The data for this work originates from an interrupted time series of nationwide cross-sectional studies. This methodology was chosen on the premise that valid inferences of change in population values can be made on the basis of repeated cross-sections within the single population [19].

The listing of the Estonian Population Registry [20] was used as a sampling frame, and all individuals aged 18 years and older were eligible for study participantion.

Using standardised methodology (population-based, random stratified sampling) 11 cross-sectional studies were conducted with data collection during April 23-29, April 30 – May 6, May 22-31, June 11-22, August 6-25, September 21 – October 3, November 11-19, November 26 – December 6, December 11-20 in 2020, and January 7-18, January 21 – February 2 in 2021. For each study, multistage stratified random sampling was used. Primary sampling strata consisted of all counties (*n*=15) and two most populated cities were considered separately from their respective counties. In each primary sampling strata, stratified by gender and age (18-39, 40-64 and 65+ years), random samples (n=200 in most regions, *n*=400 in the three most populated areas) of civilian residents were recruited.

#### Sample size

The required total sample size for individual SARS-CoV-2 RNA testing studies was estimated based on the upper Clopper-Pearson confidence limit under the assumption of no positive test results. The sample size of 2,000 was derived at a 5% level of significance with an upper confidence limit of 0.184%.

#### Study procedures

Participants were contacted by e-mail (original invitation and up to two reminders) or telephone (for those aged 65 years or older) for completion of a screening questionnaire regarding previous SARS-CoV-2 testing and symptoms of COVID-19. Respondents could take a phone interview in case of any problems with accessing the web questionnaire. A structured questionnaire (based on instrument recommended by WHO [21]) was used to elicit respondent sociodemographic data, data on the size and age structure of the household, health status, social and work related contacts within two weeks before the study.

Referral and registration for SARS-CoV-2 testing at state drive-in sites or home visit by the testing station team (for those study participants unable to access drive-in stations) was undertaken by the study team.

#### SARS-Cov-2 testing

The nasopharyngeal samples collected were tested for SARS-CoV-2 RNA by quantitative reverse-transcriptase–polymerase-chain-reaction (RT-PCR) at the SYNLAB Laboratory, a private medical laboratory company (SolGent DiaPlexQT Novel Coronavirus (2019-nCoV) Detection Kit CE-IVD). Viral RNA from all samples were isolated within 24 hours.

All SARS-CoV-2 test results were entered into the state E-Health service system and communicated back to participants by the authorised staff member of the testing stations. Participants who tested positive for SARS-CoV-2 were required to self-isolate for 14 days since developing symptoms. All those who tested positive were monitored by their own family doctor until recovery.

### Statistical analysis

Descriptive statistics (i.e. proportions and means) are presented. SARS-CoV-2 prevalence (the proportion of testing positive) and 95% Clopper-Pearson confidence interval were calculated, taking into account the sample design. Prevalence rates were calculated using the Estonian population at the beginning of 2020 as a denominator [17].

A survey-adjusted logistic regression model was applied to explore associations between data collection timing (study round), age, gender, preferred language, region of residence, size and age structure of the household, pre-existing physician diagnosed chronic conditions, body mass index, number of contacts within two weeks before the study, and having COVID-19 specific symptoms at the time of study) with the SARS-CoV-2 RNA test positivity. Variables identified as statistically significant predictors with a significance level of p < 0.05 were inserted into a multivariable logistic model (Supplement, Table 2).

We present adjusted odds ratios (OR) together with the 95% confidents estimates. Since the observed prevalence is relatively low, the odds ratios found in the logistic regression model approximate the risk ratios reasonably well.

Besides the logistic regression (i.e. binary linear model with logit-link), models with probit, complementary log-log and Cauchit link were also found. Obtained logistic regression model was the best among these competitors based on Akaike information criterion (AIC), with complementary log-log link being a close second.

We used the R statistical programming language for the analyses [22].

### Patient and Public Involvement

There were no patients or members of the public involved in the design, implementation, analyses or reporting of our research.

## Results

### SARS-CoV-2 community prevalence over the first year of the epidemic

A total of 34,915 individuals, including 15,203 males and 19,712 females participated in the series of cross-sectional studies from April 2020 to February 2021. The age of the study participants ranged from 18 to 96 years (average age 48.1 years), 85.5% filled the survey in Estonian and 14.2% in Russian language. Average household size among the study participants was 2.7. SARS-CoV-2 prevalence declined at the beginning of the observation period (in April: 0.27%, 95% CI 0.10% - 0.59%; June 2020: 0.00%, 95% CI 0.00% - 0.12%)) and remained low until the end of September 2020 (0.01%, 95% CI 0.00% - 0.17%). Since then, SARS-CoV-2 positivity rates have been increasing from 0.22% (95% CI 0.08% - 0.49%) in September to 1.27% (95% CI 0.18% - 0.68%) in November 2020, and then reaching an all-time high at 2.69% (95% CI 2.08% - 2.69%) by mid-January, 2021. About 34% of individuals (*n*= 11,879) self-reported experiencing COVID-19 symptoms at study participation (34.8% of participants testing negative, and 52.1% testing positive). Of all people tested for SARS-CoV-2, 190 were RNA-positive (see Supplement, Table 1).

Modelling confirms significant changes in SARS-CoV-2 prevalence over the first year of the epidemic. In comparison to the first survey round (April 23-29, 2020) SARS-CoV-2 RNA prevalence was significantly lower in rounds four (June 11 - 22: OR 0.00, 95% CI 0.00-0.00) and five (August 6-25: OR 0.02, 95% CI 0.00-0.22), and started to increase from round 8 (Nov 26 – Dec 6, 2020: OR 5.35, 95% CI 1.25-22.9) onwards to round 11 (Jan 21 – Feb 2, 2021: OR 8.48, 95% CI 2.03-35.4). Furthermore, regions of the country (Ida-Viru county OR 3.05, 95% CI 1.67-5.59), increasing number of household members (for one additional OR 1.15, 95% CI 1.02-1.29), reporting symptoms of COVID-19 (OR 2.21, 95% CI 1.59-3.09), and completion of the survey in Russian (OR 1.85, 95% CI 1.15-2.99) were all associated with higher SARS-CoV-2 RNA positivity (see Supplement, Table 2).

## Discussion

The nationwide study documents substantial changes in the population prevalence of SARS-CoV-2 RNA in Estonia during the 1^st^ year of the COVID-19 epidemic, with an initial decrease between April and June, 2020. The findings of the post 1^st^ wave of COVID-19 prevalence and decline are in perfect agreement with a community-based SARS-CoV-2 study from England for the period of April to June 2020 [9]. In their study, SARS-CoV-2 community prevalence of 0.32%, (95% CrI 0.19%-0.52%) in April 2020 declined to a very low level by the end of June 2020 (0.08%, 95% CrI 0.05%-0.12%). In Estonia, the short period of very low SARS-CoV-2 prevalence over the summer of 2020 was followed by an initially slow (in September and October) and then escalating increase since November 2020.

This study documents a clear decline in the prevalence of SARS-CoV-2 following the implementation of the nationwide non-pharmacological intervention (NPI) at the beginning of the epidemic. SARS-CoV-2 prevalence remained extremely low for a short period after lifting NPI measures. In the face of mitigation (slowing down transmission) rather than suppression (stopping SARS-CoV-2 community spread) of containment, an exponential increase of new COVID-19 cases occurred at the verge of the 2^nd^ year of the epidemic.

These findings allow us to speculate that, until now, this is a very unforgiving virus. While rigorous and comprehensive NPI measures are clearly effective in stopping transmission, lifting the measures or less stringent implementation will lead to new and sizable outbreaks.

Second, findings from Estonia should be interpreted in the context of the high SARS-CoV-2 testing rate (80,630/100,000),[23] a very low COVID-19 case fatality rate of 0.8% (both, as of March 18, 2021) and no significant excess (all cause) deaths over the first year of the epidemic [24].

We saw that those with a larger household size were at higher risk of SARS-CoV-2 infection with no attributable risk either from the age of the individual or the age structure of the household (very similar to the results of the study from UK [25]). Ongoing household transmission with occasional spill over to other households could act as an important driver for ongoing transmission,[26] and is estimated to be responsible for roughly 70% of SARS-CoV-2 transmission when widespread community control measures are in place [27].

Our findings of higher SARS-CoV-2 risk among those reporting symptoms characteristic to COVID-19 are clearly not new. Yet, it highlights the need to focus on symptomatic cases rather than mass-testing in the face of resource constraints or competing resource needs (i.e. vaccination). Focus on symptomatic COVID-19 cases has a solid evidence base - the majority of COVID-19 cases are symptomatic (∼ 60-80%) [28], and are significantly more likely to infect their close contacts than their asymptomatic counterparts [29].

Last but not least, we saw regional and ethnic (main language spoken) differences in SARS-CoV-2 positivity. Disproportionately affected racial and ethnic minority groups have been reported elsewhere (United States,[30] UK, [31]). In Estonia, ethnic disparities are not unique to COVID-19 outcomes [32,]. The reasons for ethnic disparities in COVID-19 outcomes are multi-layered,[31] and underline the regional differences in Estonia. Ida-Viru County is in the north-eastern part of Estonia bordering the Russian Federation. The overwhelming majority (82%) of residents are Russian speaking. It is important to note that nearly 75% of Russian speakers in Estonia regularly follow TV channels and online media originating from the Russian Federation,[33] and are more likely to trust Russian than domestic [Estonian] or EU media.[34] Whether the Russian Federation’s pandemic-related disinformation campaign [35] has had some effect on the beliefs and behaviours of the Russian speaking population in Estonia (and other neighbouring countries with sizable Russian speaking minorities) is unknown at this stage. There are anecdotal reports from Ida-Viru County on residents of declining state-provided COVID-19 vaccines and demands to be vaccinated with the Russian Sputnic vaccine.[36] There is a risk that COVID-19 vaccine uptake will be lower among minority ethnic groups in Estonia, thereby widening the health gap further. COVID-19 risk communication and community engagement is a priority for information provision and to counter misinformation.

In conclusion, a rather limited number of studies have assessed the prevalence of SARS-CoV-2 infection in the general population (seroprevalence,[37,38] SARS-CoV-2 RNA [10,39,40]). Population-based studies assessing temporal changes in SARS-CoV-2 prevalence, either via repeated cross-sectional studies [41] or following subjects longitudinally,[9] are, to our best knowledge, exceedingly rare. It is critically important to create a knowledge base to inform future strategies, and a range of real-life COVID-19 epidemic scenarios over extended periods needs to be documented to assist in understanding the infection risk factors at the individual and population levels. Analyses based on patients in need of hospital treatment, and/or with co-morbidities reported during the early phases of the COVID-19 epidemic, were unable to disentangle infection from virulence risks. Yet, primary prevention operates through the control of (the true) infection risk factors.

Our study has several limitations. The degree to which the study is representative of the larger population is influenced by the low response rate and potential selective factors associated with responses. To minimise non-response bias, the prevalence estimates were weighted (age, gender, and region) to ensure representativeness of the source population. Yet, there could be other factors for which we did not have detailed information about population distributions which are also associated with testing positive for SARS-CoV-2. The number of people testing SARS-CoV-2 RNA positive in the cross-sectional studies is low leading to relatively large uncertainty around estimates.

We see the long period of observation and population-based nationwide study design as strengths of our work. Interpretation of changes in SARS-CoV-2 incidence and positivity rates originating from case notification or clinical cases is likely to be confounded by substantial changes in testing practice over time. Our study is based on a series of cross-sectional studies with a standardised methodology, and is thereby very unlikely to be influenced by the testing practice. As this evaluation is based upon observing a single population over time, we speculate that selection bias or unmeasured confounders would operate rather uniformly over the period of observation, though presenting a less threatening trend of SARS-CoV-2 prevalence and analysis of factors associated with SARS-CoV-2 positivity.

## Conclusions

Focusing only on medical interventions (testing, treatment, vaccination) for epidemic control can create false public expectations of a return to, or maintenance of, normality. The population-based effect of the novel vaccines against SARS-CoV-2 is highly contingent on the infection-blocking (or transmission-blocking) action of the vaccine and population uptake [8]. SARS-CoV-2 population prevalence needs to be carefully monitored to inform containment decisions as vaccine programmes are rolled out.

## Data Availability

Aggregated level data on COVID-19 is publicly available (open data at https://koroona.ut.ee, and at https://koroonakaart.ee/en). Data are available upon reasonable request from Mikk Jurisson.

## Supplementary materials

Table 1. Characteristics of the population-based SARS-CoV-2 prevalence studies and respective study participants, Estonia, 2020-2021.

Table 2. Risk factors for testing positive for SARS-CoV-2, population-based SARS-CoV-2 prevalence studies, Estonia, 2020-2021.

## Contributorship statement

Anneli Uusküla - Conceptualisation, Methodology, Writing - Original Draft; Ruth Kalda, Mihkel Solvak, Mikk Jürisson, Krista Fischer, Aime Keis, Kristjan Vassil, Jaak Vilo, Hedi Peterson, Lili Milani, Mait Metspalu - Conceptualisation, Writing - Review & Editing; Meelis Käärik, Krista Fischer, Uku Raudvere, Ene Käärik, Liis Kolberg, Tuuli Jürgenson, Ene-Margit Tiit - Investigation, Data analysis, Writing - Review & Editing; Meelis Käärik, Krista Fischer - Visualisation. All listed authors reviewed and edited the manuscript and approved the final, submitted version. All authors confirm that they had full access to the data in the study and accept responsibility to submit for publication.

## Data availability statement

Aggregated level data on COVID-19 is publicly available (open data at https://koroona.ut.ee, and at https://koroonakaart.ee/en). Data are available upon reasonable request from Mikk Jürisson.

## Acknowledgments

This study was funded by grants SMVPT20243, SMVPT20599 from the Estonian Ministry of Education and Research; Estonian Research Council Grants (PRG1095, PRG59, IUT34-4), and (Project No 2014-2020.4.01.16-0271, ELIXIR).

We thank our study partners Kantar-Emor AS, OÜ Medicum Eriarstiabi, and SYNLAB Eesti OÜ.

## Ethics approval

Ethical approval for the study was obtained from the Research Ethics Committee of the University of Tartu.

