## Supplemental Tables for "The 1st year of the COVID-19 epidemic in Estonia: an interrupted time series of population based nationwide cross-sectional studies"

**Appendix**

**Table 1. Characteristics of the population-based SARS-CoV-2 prevalence studies and respective study participants, Estonia, 2020-2021.**

| | Round 1 | Round 2 | Round 3 | Round 4 | Round 5 | Round 6 | Round 7 | Round 8 | Round 9 | Round 10 | Round 11 |
|---|---|---|---|---|---|---|---|---|---|---|---|
| | April 23–29, 2020 | April 30–May 6, 2020 | May 22–31, 2020 | June 11–22, 2020 | Aug 6–25, 2020 | Sept 21–Oct 3, 2020 | Nov 11–19, 2020 | Nov 26–Dec 6, 2020 | Dec 11–20, 2020 | Jan 7–18, 2021 | Jan 21–Feb 2, 2021 |
| *Study characteristics* | | | | | | | | | | | |
| Total sample | 10 209 | 12 020 | 21 830 | 28 034 | 25 998 | 22 900 | 23 187 | 20 032 | 23 921 | 21 063 | 25 135 |
| Non-contacts (*n*) | 4 119 | 6 113 | 12 869 | 20 133 | 19 467 | 15 460 | 16 322 | 14 296 | 17 900 | 14 957 | 18 042 |
| Refusals (*n*) | 2 060 | 1 791 | 2 923 | 2 414 | 1 546 | 3 024 | 2 623 | 2 211 | 2 294 | 2 583 | 2 816 |
| Other non-response (*n*) | 1 141 | 981 | 2 538 | 1 615 | 1 813 | 983 | 893 | 688 | 749 | 732 | 1 318 |
| Participants (*n*) | 2 889 | 3 135 | 3 500 | 3 872 | 3 172 | 3 433 | 3 349 | 2 837 | 2 978 | 2 791 | 2 959 |
| SARS-CoV-2 tested (*n*) | 2 306 | 2 666 | 2 579 | 2 983 | 2 335 | 2 532 | 2 726 | 2 381 | 2 522 | 2 370 | 2 470 |
| *Participants characteristics* | | | | | | | | | | | |
| Men (*n*, %) | 1254, 43.4% | 1377, 43.9% | 1627, 46.5% | 1657, 42.8% | 1413, 44.6% | 1504, 43.8% | 1388, 41,5% | 1189, 41,9% | 1297, 43,6% | 1202, 43,1% | 1295, 43,8% |
| Age (mean, SD, range) | 47.7, 15.8, 18-94 | 46.7, 15.6, 18-94 | 49.6, 16.7, 18-93 | 47.5, 15.6, 18-92 | 47.2, 15.9, 18-94 | 48.2, 15.8, 18-95 | 48.7, 15.9, 18-96 | 49.9, 16.2, 18-94 | 47.0, 15.8, 18-91 | 48.1, 16.1, 18-93 | 48.6, 16.0, 18-93 |
| Size of the household (mean, SD) | 2.77, 1.42 | 2.79, 1.42 | 2.70, 1.41 | 2.75, 1.41 | 2.77, 1.43 | 2.68, 1.36 | 2.62, 1.36 | 2.63, 1.36 | 2.68, 1.37 | 2.66, 1.36 | 2.67, 1.38 |
| Respondent language Russian (yes; *n*, %) | 396, 13.7% | 432, 13.8% | 513, 14.7% | 554, 14.3% | 380, 12.0% | 482, 14.0% | 565, 16.9% | 411, 14.5% | 388, 13.0% | 406, 14.6% | 428, 14.5% |
| Smoking (yes; *n*, %) | 685, 23.7% | 709, 22.6% | 762, 21.8% | 789, 20.4% | 706, 22.3% | 681, 19.9% | 652, 19.5% | 520, 18.3% | 600, 20.2% | 555, 19.9% | 553, 18.7% |
| Pre-existing chronic disease (yes; *n*, %) | 1138, 39.4% | 1217, 38.8% | 1493, 42.7% | 1532, 39.6% | 1224, 38.6% | 1375, 40.1% | 1345, 40.2% | 1176, 41.2% | 1120, 37.6% | 1123, 40.2% | 1192, 40.3% |

| | | | | | | | | | | | |
|---|---|---|---|---|---|---|---|---|---|---|---|
| Self-reported COVID-19 symptoms[1] (yes; *n*, %) | 1079, 37.4% | 1132, 36.1% | 1110, 31.7% | 1142, 29.5% | 1047, 33.0% | 1234, 36.0% | 1159, 34.6% | 1005, 35.4% | 1987, 36.5% | 939, 33.6% | 945, 31.9% |
| Previous SARS-CoV-2 testing (yes; *n*, %) | 143, 4.95% | 159, 5.07% | 318, 9.09% | 308, 7.95% | 363, 11.4% | 810, 23.6% | 1107, 33.1% | 1061, 37.4% | 1268, 42.6% | 1341, 48.1% | 1475, 49.9% |
| Previously tested positive for SARS-CoV-2 RNA (*n*, %) | 12, 8.39% | 11, 6.92% | 16, 5.03% | 15, 4.87% | 6, 1.65% | 14, 1.73% | 19, 1.72% | 31, 2.92% | 43, 3.39% | 75, 5.59% | 82, 5.56% |
| *SARS-CoV-2 positivity and estimated prevalence* | | | | | | | | | | | |
| No of test positives | 4 | 8 | 2 | 0 | 1 | 5 | 10 | 30 | 31 | 55 | 42 |
| Prevalence (%, 95% CI) | 0.27% (0.10% - 0.59%) | 0.17% (0.05% - 0.41%) | 0.04% (0.00% - 0.22%) | 0.00% (0.00% - 0.12%) | 0.01% (0.00% - 0.17%) | 0.22% (0.08% - 0.49%) | 0.37% (0.18% - 0.68%) | 1.34% (0.92% - 1.89%) | 1.27% (0.87% - 1.79%) | 2.69% (2.08% - 2.69%) | 2.05% (1.53% - 2.69%) |

**Table 2. Risk factors for testing positive for SARS-CoV-2, population-based SARS-CoV-2 prevalence studies, Estonia, 2020-2021.**

| **Variables** | **Odds ratio (OR)** | **Lower confidence limit (2.5%)** | **Upper confidence limit (97.5%)** | **p-value [1]** |
|---|---|---|---|---|
| *Data collection timing* | | | | |
| April 23–29, 2020 (base) | 1 | | | |
| April 30–May 6, 2020 | 0,63 | 0,13 | 3,10 | |
| May 22–31, 2020 | 0,67 | 0,08 | 5,39 | |
| June 11–22, 2020 | 0,00 | 0,00 | 0,00 | *** |
| Aug 6–25, 2020 | 0,02 | 0,00 | 0,22 | ** |
| Sept 21–Oct 3, 2020 | 0,84 | 0,16 | 4,43 | |
| Nov 11–19, 2020 | 1,44 | 0,30 | 6,84 | |
| Nov 26–Dec 6, 2020 | 5,35 | 1,25 | 22,93 | * |
| Dec 11–20, 2020 | 5,12 | 1,21 | 21,73 | * |
| Jan 7–18, 2021 | 11,07 | 2,65 | 46,32 | *** |
| Jan 21– Feb 2, 2021 | 8,48 | 2,03 | 35,43 | ** |
| *Participant Language* | | | | |
| Estonian (base) | 1,00 | | | |
| Russian | 1,85 | 1,15 | 2,99 | * |
| *Size of the household (number of individuals)* | 1,15 | 1,02 | 1,29 | * |
| *Reporting symptoms[2] at the time of study* | | | | |
| No | 1,00 | | | |
| Yes | 2,21 | 1,59 | 3,08 | *** |
| *Region of the country* | | | | |
| Harju county w/o Tallinn (base) | 1,00 | | | |
| Hiiu county | 0,91 | 0,32 | 2,60 | |

| | | | | |
|---|---|---|---|---|
| Ida-Viru county | 3,06 | 1,67 | 5,59 | *** |
| Jõgeva county | 0,14 | 0,02 | 1,03 | |
| Järva county | 0,54 | 0,14 | 2,05 | |
| Lääne-Viru county | 1,08 | 0,38 | 3,04 | |
| Lääne county | 0,24 | 0,05 | 1,06 | |
| Põlva county | 0,13 | 0,02 | 1,00 | |
| Pärnu county | 0,86 | 0,37 | 1,99 | |
| Rapla county | 0,38 | 0,11 | 1,32 | |
| Saare county | 0,93 | 0,38 | 2,27 | |
| Tallinn city | 1,32 | 0,74 | 2,34 | |
| Tartu city | 0,87 | 0,36 | 2,11 | |
| Tartu county w/o city | 0,50 | 0,17 | 1,52 | |
| Valga county | 0,83 | 0,19 | 3,60 | |
| Viljandi county | 2,20 | 0,95 | 5,12 | . |
| Võru county | 1,71 | 0,74 | 3,91 | |

[1] *** $p< 0.001$; ** $p< 0.01$; * $p< 0.05$

[2] Participants reporting at least one of the three major symptoms (cough, fever, dyspnea) or at least two of minor symptoms (fatigue, sputum production, muscle or joint aches, headache, new loss of taste or smell, sore throat, congestion or runny nose, nausea or vomiting, diarrhea, irritability or confusion)
